# Ex-PRESS implantation versus trabeculectomy for long-term maintenance of low intraocular pressure in patients with open angle glaucoma

**DOI:** 10.1101/2022.09.10.22279798

**Authors:** Kana Tokumo, Naoki Okada, Hiromitsu Onoe, Kaori Komatsu, Shun Masuda, Hideaki Okumichi, Kazuyuki Hirooka, Ryo Asaoka, Yoshiaki Kiuchi

## Abstract

**Purpose:** To compare the efficacy of Ex-PRESS implantation to trabeculectomy with mitomycin C, for maintaining low target intraocular pressure (IOP) in patients with open angle glaucoma.

**Materials and Methods:** Patients were randomly assigned to receive Ex-PRESS implantation or trabeculectomy. Patients with IOP ≥ 15 mmHg were included in this study. Surgical success was defined according to three target mean IOP ranges (5 mmHg ≤ IOP ≤18 mmHg [criterion A], 5 mmHg ≤ IOP ≤15 mmHg [criterion B], and 5 mmHg ≤ IOP ≤12 mmHg [criterion C]) representing reductions of at least 20% below baseline on two consecutive follow-up visits 3 months post surgery.

**Results:** A total of 73 patients, including 30 in the Ex-PRESS implantation group and 43 in the trabeculectomy group, were included in the study. The baseline IOP was 20.4 ± 4.9 mmHg in the Ex-PRESS implantation group and 21.9 ± 7.9 mmHg in the trabeculectomy group. There were no significant differences in baseline ocular or demographic characteristics between the two groups. There was no statistical difference in IOP every 6 months. After the 3-year follow-up, success rates were A) 60.0% and 60.2%, B) 45.7% and 58.1%, and C) 31.5% and 40.5% for the Ex-PRESS implantation and trabeculectomy groups, respectively. A greater number of glaucoma medications before surgery was associated with a higher failure rate in the trabeculectomy group but not the Ex-PRESS implantation group.

**Conclusions:** Both procedures resulted in similar IOP reductions and success rates for low target IOP. The number of preoperative glaucoma medications was a risk factor for trabeculectomy failure.

## Introduction

Filtering surgery is widely used for the treatment of glaucoma with uncontrollable intraocular pressure (IOP). Currently, the most popular filtering surgery is trabeculectomy (TLE), in which aqueous humor filtration is enhanced and IOP reduced by creating a scleral flap that leads to bleb formation in the conjunctiva. The success and complication rates for TLE are well established [1-2].

The Ex-PRESS glaucoma filtration device (Alcon Laboratories, Fort Worth, TX, USA) was introduced as a modification to TLE. Although the surgical procedure and postoperative management is similar to TLE, Ex-PRESS (EXP) surgery does not require sclerectomy or iridectomy. Therefore, postoperative inflammation and hemorrhage may be reduced. Both surgical procedures can achieve similar IOP reductions with few complications when the target pressure is set to 18 mmHg [3-5]. However, the optimal target IOP for glaucoma treatment depends on disease stage, IOP before treatment, age, and other risk factors. In general, more severe glaucoma requires lower IOP post surgery to maintain visual function [6-8].

The success of TLE depends on preoperative IOP, previous surgery, number of preoperative medications, and race [9]. There is a high incidence of normal tension glaucoma (NTG) in the Japanese population [10], and visual field impairment in subjects with NTG often progresses even when IOP is within the normal range. Therefore, NTG patients may benefit from a postoperative target pressure below that of patients with higher preoperative IOP.

This study compared the efficacy and safety of EXP to TLE for achieving target postoperative IOPs of ≤18, ≤15, and ≤12 mmHg, each representing reductions of at least 20% below baseline, among patients in the Japanese population with open angle glaucoma (OAG, including exfoliation glaucoma and primary OAG).

## Materials and methods

The institutional review board of Hiroshima University approved the study protocol before recruitment. This study was registered with the University Hospital Medical Information Network Clinical Trials Registry of Japan (identifier University Hospital Medical Information Network 000008981; date of access and registration, September 25, 2012). All patients provided written informed consent before participation, and the study was conducted in accordance with the tenets of the Declaration of Helsinki.

### Randomization and treatment

The study was conducted at the Department of Ophthalmology, Hiroshima University Hospital. Patients were enrolled between November 2012 and March 2015. Follow-up was done for one year, three years if possible. Patients were randomized to receive TLE or EXP using a computer-based random number generator. Clinicians enrolled participants to this study. Random allocation were prepared by staff not associated with this study and he assigned participants to interventions. It was confirmed after patient enrollment is completed which surgery will be performed. Both patients and clinicians were aware of the therapy (therefore, this was an open-label, randomized study).

### Patients and inclusion criteria

This was a single-center clinical trial of OAG patients > 20 years and older with or without cataract scheduled for filtration surgery. Patients who previously underwent glaucoma surgery were excluded. However, subjects who underwent cataract surgery more than 6 months prior to planned TLE or EXP were included. We analyzed the first eye when both eyes met the inclusion criteria. Subjects with preoperative IOP < 15 mmHg were excluded after randomization, and their data were analyzed in another study.

### Outcome measures

Goldman applanation tonometry was used to measure the IOP. Preoperative IOP and best-corrected visual acuity (VA) were recorded during the examination immediately before surgery. We documented the IOP and VA the day after surgery and at every follow-up visit. Postoperative IOP and VA values measured every six months for three years were included in the efficacy (surgical success) analysis. The surgical procedures and postoperative management were similar for each case.

Anesthesia included subconjunctival 2% lidocaine injection. A limbal-based conjunctival flap was utilized, followed by the creation of a half-thickness scleral flap. Small pieces of gelatin sponge soaked with 0.4 mg/mL mitomycin C (MMC) was applied to the exposed tissue, including the posterior surface of Tenon’s capsule and conjunctiva, adjacent episcleral tissue, and scleral flap for 5 min. After 5 min, all sponges were removed, and the wound was irrigated with 100 mL of balanced salt solution. For patients assigned TLE, a block incision was made in the region of the trabecular meshwork, followed by peripheral iridectomy. For those patients randomized to receive EXP, a small scleral tunnel was created as a shunt in the region of the trabecular meshwork using a 25-G needle. The scleral flap and conjunctiva were closed with 10-0 nylon sutures.

Patients were treated with laser suture lysis if IOP was higher than the target IOP. The target IOP for most patients was set at 8–10 mmHg 2 weeks postoperatively.^8^

All patients received a similar topical medical regimen, including postoperatively: 1.5% levofloxacin and 0.1% topical betamethasone three times per day. Patients receiving combined cataract surgery were also administered nepafenac three times a day. Topical atropine sulfate (1%) was used as needed. All doses were tapered at the surgeon’s discretion.

### Parameters for surgical success

The primary outcome was surgical success based on target postoperative IOP range at 6-month intervals with or without antiglaucoma medications according to guidelines by the World Glaucoma Association [11]. There were three target IOP ranges: (A) 5 mmHg ≤IOP ≤18 mmHg, (B) 5 mmHg ≤IOP ≤15 mmHg, and (C) 5 mmHg ≤IOP ≤12 mmHg. Failure was defined as an IOP above the indicated range on two consecutive follow-up visits 3 months after surgery and a reduction of less than 20% below baseline, or the need for further glaucoma surgery. The need for laser suture lysis or bleb needling was not considered surgical failure as these procedures are part of routine postoperative management for filtration surgeries.

### Statistical analysis

All statistical analyses were performed using JMP version 15 (Cary, North Carolina, USA). Results are presented as mean ± standard deviation (SD) unless otherwise indicated. Mean values of normally distributed continuous variables were compared between groups by Student’s t-test and proportion (e.g., of patients with visual stability or surgical success) by chi-square test. Measured IOP values were compared using the Mann–Whitney nonparametric test with Bonferroni correction for repeated measures. Patients requiring additional surgery were censored from the analysis after reoperation. Success rates were analyzed over time using Kaplan–Meier survival curves. The survival data were analyzed using regression analysis based on the Cox proportional hazards model. All statistical tests were two-sided, and P-values <0.05 were considered statistically significant. A sample size calculation determined that 30 eyes in each group were required to detect a 2.5-mmHg difference in IOP with an SD of 2.75 mmHg and power of 80%. To account for patient dropout, 73 eyes were included.

## Results

A total of 105 patients with OAG (105 eyes) were enrolled in the study. Two patients who had glaucoma surgery were excluded. After randomization, 30 patients whose preoperative IOP was <15 mmHg were excluded. Thirty eyes were treated with EX-PRESS implantation (EXP group) and 43 eyes with TLE (TLE group). The number of patients who could be followed is shown in Fig 1. Patients who had mild to severe cataract underwent combined cataract surgery. All patients were Japanese. Patient demographics and basic characteristics are summarized in Table 1. The mean follow-up period was 25.9 ± 12.9 months in the EXP group and 23.8 ± 13.1 months in the TLE group. No differences were found in baseline IOP, sex ratio, age, and number of glaucoma medications between EXP and TLE groups.

**Table 1.** Baseline demographics and ocular characteristics of the EXP group (30 eyes) and TLE group (43 eyes). EXP, Ex-PRESS; IOP, intraocular pressure; LogMAR, logarithm of the minimal angle of resolution; SD, standard deviation; TLE, trabeculectomy *Student’s t-tests ‡Mann–Whitney nonparametric tests †Pearson chi-square tests

| Characteristics | EXP group (n = 30) | TLE group (n = 43) | P-values |
| --- | --- | --- | --- |
| Age (years), mean ± SD | 68.3 ± 11.9 | 68.7 ± 12.4 | 0.90* |
| Sex, n (%) |  |  | 0.64<sup>†</sup> |
| Male | 17 (56.7) | 22 (51.2) |  |
| Female | 13 (43.3) | 21 (48.8) |  |
| IOP (mmHg), mean ± SD | 20.4 ± 4.9 | 21.9 ± 7.9 | 0.84<sup>‡</sup> |
| Glaucoma medications, n, mean ± SD | 3.1 ± 0.8 | 3.3 ± 0.8 | 0.44* |
| Follow-up period (months), mean ± SD | 25.9 ± 12.9 | 23.8 ± 13.1 | 0.50* |
| Previous cataract surgery, n (%) | 11(36.7) | 11 (19.6) | 0.31* |
| Surgery (%) |  |  | 0.59<sup>†</sup> |
| Glaucoma surgery alone | 22 (73.3) | 29 (67.4) |  |
| Combined glaucoma and cataract surgery | 8 (26.7) | 14 (32.6) |  |

**Fig 1.**
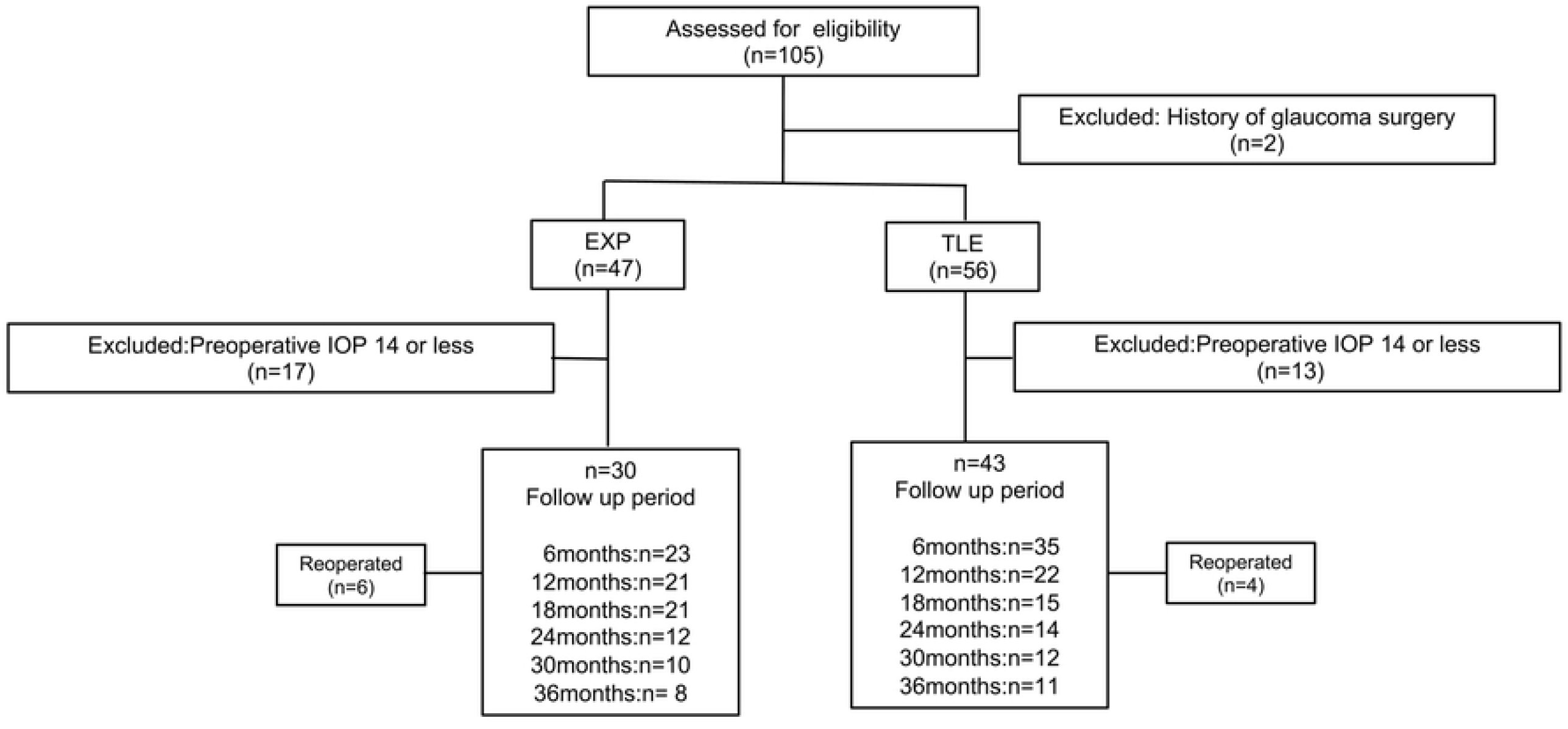
Flowchart of follow-up.

### IOP reduction

The mean IOP at each postoperative visit is presented in Table 2 and shown graphically in Fig 2. Both surgical procedures led to a significant sustained reduction in IOP. In the EXP group, the IOP decreased from 20.4 ± 4.9 to 13.2 ± 1.9 mmHg at the 3-year follow-up visit (P = 0.001, paired t-tests). In the TLE group, the IOP decreased from 21.9 ± 7.9 to13.5 ± 5.0 mmHg at the 3-year follow-up visit (P = 0.0012, paired t-tests). Both surgical procedures resulted in a significant reduction in IOP after surgery compared to the preoperative IOP. There were no differences in IOP between the two groups during observation.

**Table 2.** Mean intraocular pressure and medication use.

|  | EXP group (n = 30) | TLE group (n = 43) | P-values |
| --- | --- | --- | --- |
| Baseline IOP (mmHg) | $20.4 \pm 4.9$ | $21.9 \pm 7.9$ | 0.84‡ |
| Glaucoma medications, n | $3.1 \pm 0.8$ | $3.3 \pm 0.8$ | 0.44* |
| 6 months |  |  |  |
| IOP (mmHg) | $12.0 \pm 3.9$ | $11.2 \pm 2.9$ | 0.46‡ |
| Glaucoma medications, n | $0.3 \pm 0.8$ | $0.2 \pm 0.8$ | 0.83* |
| 12 months |  |  |  |
| IOP (mmHg) | 12.1 ± 3.5 | 12.4 ± 4.3 | 0.68‡ |
| Glaucoma medications, n | 0.5 ± 1.2 | 0.6 ± 1.3 | 0.82* |
| 18 months |  |  |  |
| IOP (mmHg) | 12.8 ± 3.9 | 12.8 ± 4.7 | 0.82‡ |
| Glaucoma medications, n | 1.2 ± 1.5 | 1.0 ± 1.5 | 0.70* |
| 24 months |  |  |  |
| IOP (mmHg) | 13.2 ± 3.3 | 13.1 ± 4.5 | 0.68‡ |
| Glaucoma medications, n | 1.5 ± 1.7 | 1.1 ± 1.5 | 0.52* |
| 30 months |  |  |  |
| IOP (mmHg) | 15.5 ± 5.5 | 13.4 ± 4.3 | 0.15‡ |
| Glaucoma medications, n | 1.7 ± 1.9 | 1.0 ± 1.5 | 0.29 * |
| 36 months |  |  |  |
| IOP (mmHg) | 13.2 ± 1.9 | 13.5 ± 5.0 | 0.91‡ |
| Glaucoma medications, n | 2.2 ± 1.3 | 1.2 ± 1.5 | 0.04* |

EXP, Ex-PRESS; IOP, intraocular pressure; TLE, trabeculectomy
Data are presented as mean ± SD, unless otherwise indicated.
\*Student t-tests
‡Based on Mann–Whitney nonparametric tests with Bonferroni correction, P-values <0.0083 were statistically significant.

**Fig 2.**
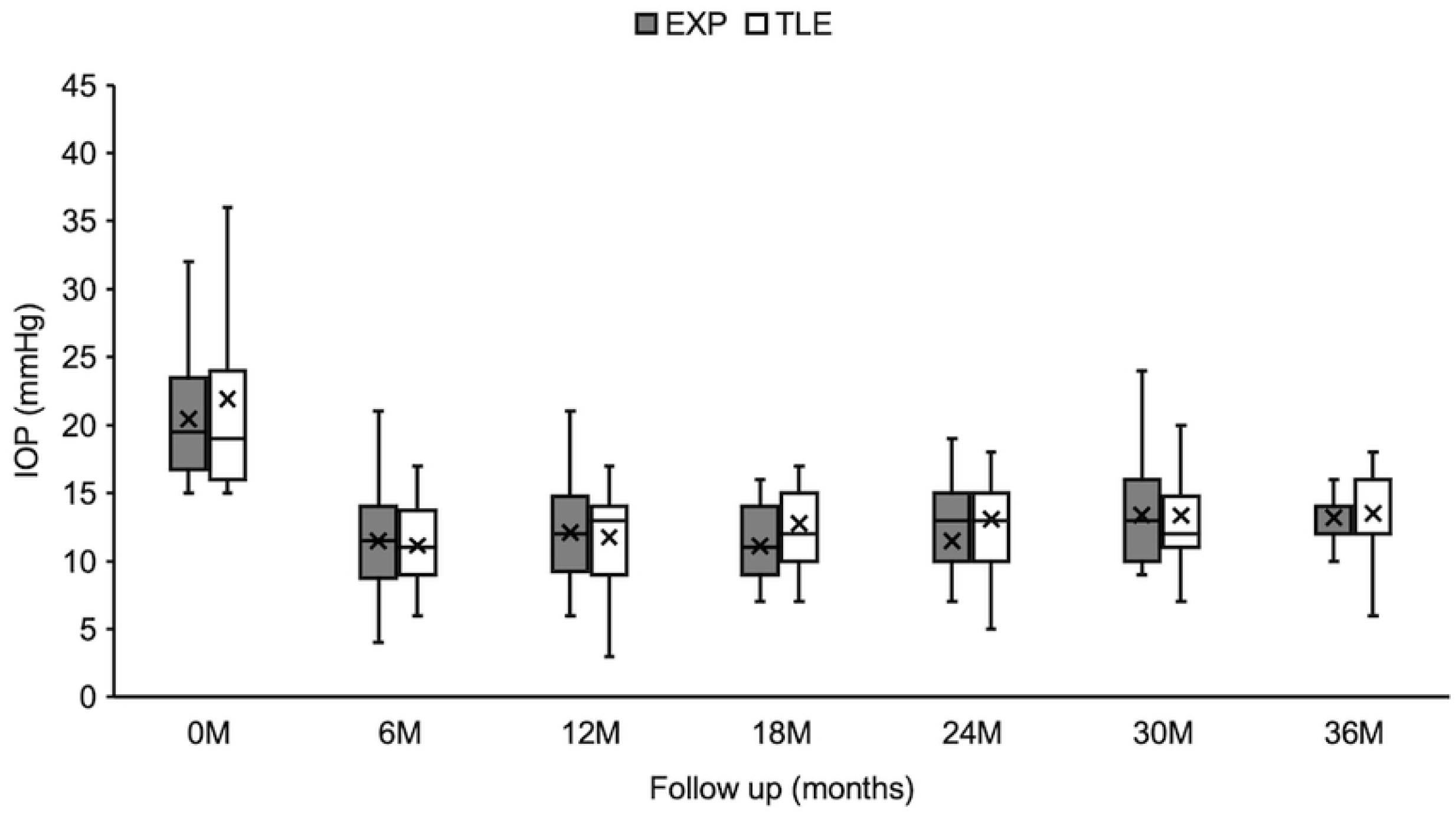
IOP (in mmHg) for the EXP (30 eyes) and TLE groups (43 eyes) from the preoperative visit to the 3-year follow-up visit (mean ± SD).

Kaplan–Meier survival analysis was used to compare success rates between the two treatment groups. We created three definitions of success and compared the survival curves. At the 1-year follow-up, the success rates of EXP and TLE were 72.0% and 70.8%, 65.3% and 68.3%, and 55.1% and 58.5% for criteria A, B, and C, respectively. At the 3-year follow-up, success rates of EXP and TLE were 60.0% and 60.2%, 45.7% and 58.1%, and 31.5%and 40.5% for criteria A, B, and C, respectively (Fig 3).

**Fig 3.**
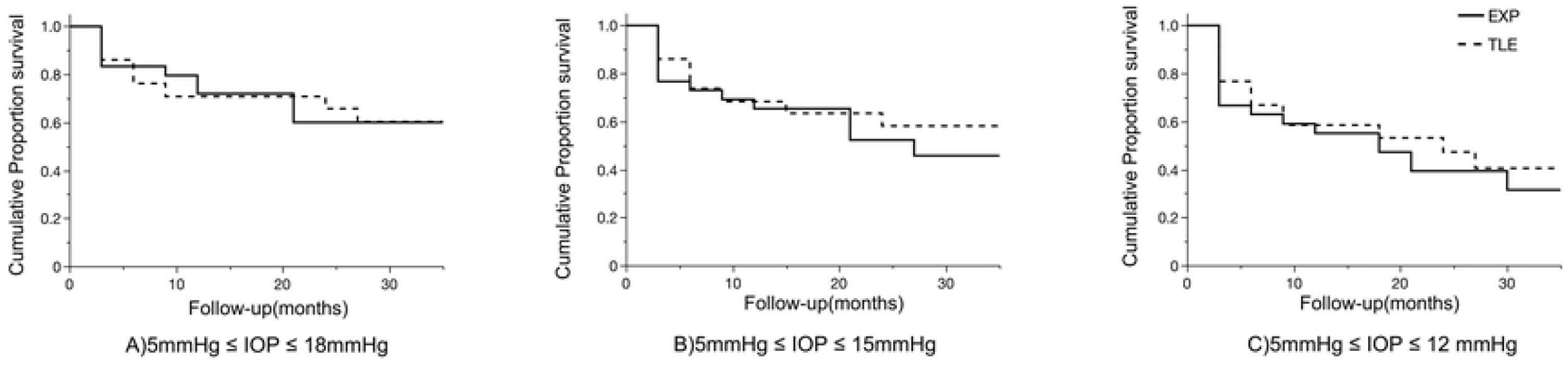
Kaplan–Meier survival curves of surgical outcomes for eyes in the EXP (30 eyes; solid line) and TLE groups (43 eyes; dotted line).

No significant differences in treatment efficacy were found between strata (A, P = 0.93; B, P = 0.53; C, P = 0.58, log-rank test).

### Glaucoma medication use

The mean number of glaucoma medications required at each postoperative visit is shown in Table 2. Both groups required significantly fewer glaucoma medications after surgery, with no significant differences between groups. The EXP group required more postoperative eye drops than the TLE group. The difference increased gradually with time.

### Visual acuities

Decimal VA was converted to logarithm of minimum angle of resolution equivalents (logMAR) for statistical analysis. All patients who underwent glaucoma surgery combined with cataract surgery showed improvement in VA, so these patients were excluded from VA analysis. Fig 4 shows the VA results 1 year after glaucoma surgery. LogMAR VA did not differ significantly between the groups immediately after the procedure (0.30 ± 0.60 in the EXP group and 0.14 ± 0.33 in TLE group, P = 0.20 by Mann–Whitney U-test), at 1 year post surgery (0.37 ± 0.61 in the EXP group and 0.20 ± 0.43 in TLE group, P = 0. 24 by Mann–Whitney U-test) or 3 years post surgery (0.44 ± 0.55 in the EXP group and 0.33 ± 0.52 in TLE group, P = 0.34 by Mann– Whitney U-test). Six patients in each group lost more than two lines at the last visit, while one subject in the TLE group developed cataract, which resulted in a decrease in VA. The others lost their vision due to glaucoma progression.

**Fig 4.**
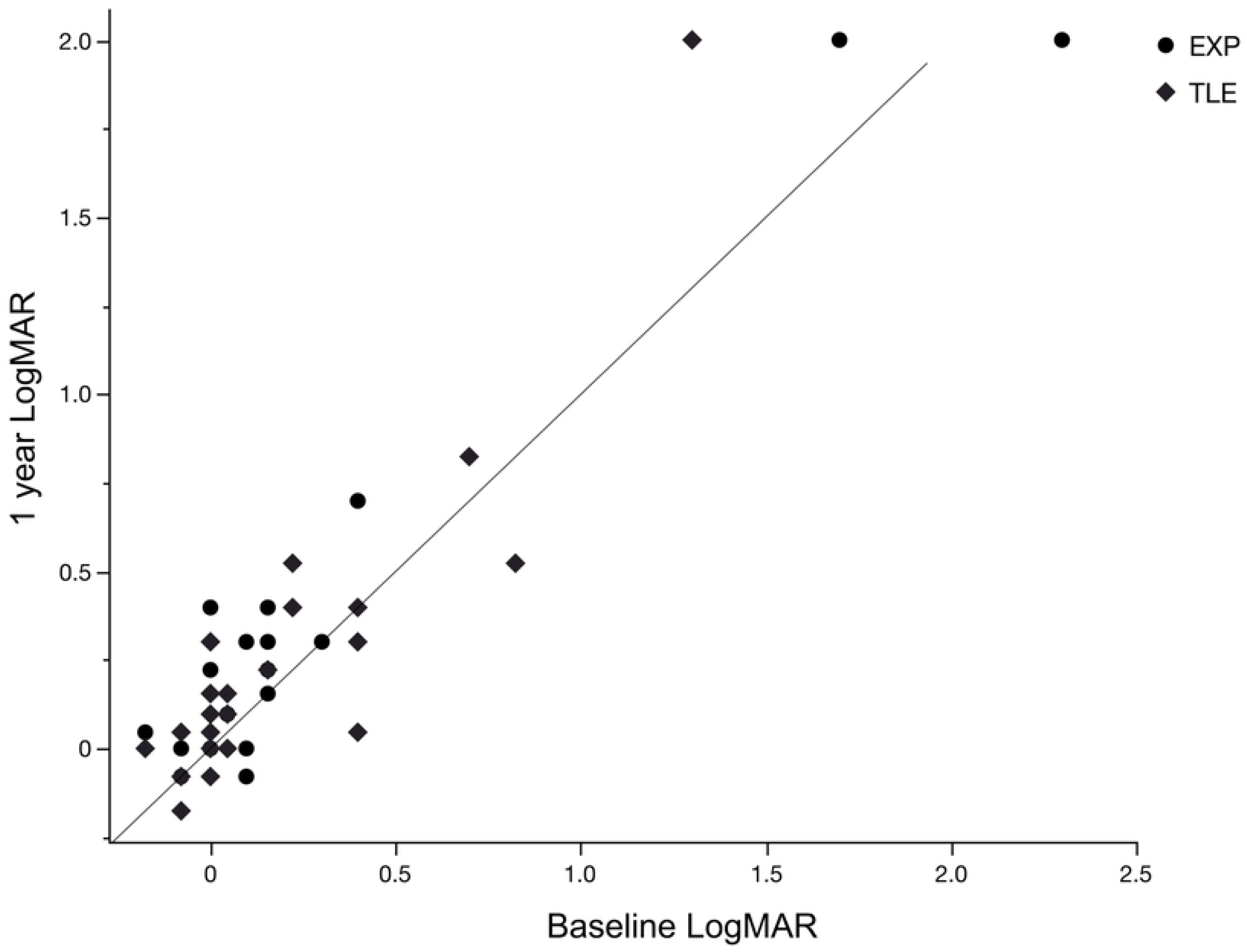
Scatterplot comparing VA (logMAR) values at baseline and 1-year visit for individual patients in the EXP (30 eyes; circular dots) and TLE groups (43 eyes; diamond dots).

### Postoperative complications

Table 3 shows complications of the two groups in the early and late postoperative periods. At the first month postoperatively, 13 patients in the EXP group and 10 patients in the TLE group developed early complications. These included hyphema, choroidal detachment, shallow anterior chamber, hypotony maculopathy, and conjunctiva leakage. Both groups showed similar complications. There was no statistically significant difference in the total prevalence of early-phase complications between groups (P = 0.07 Student t-test). Only the TLE group showed late-phase postoperative complications, one case of choroidal detachment and one of conjunctiva leakage. However, there was no statistically significant difference in the prevalence of late-phase complications between groups (P = 0.27 by Student’s t-test).

**Table 3.** Early postoperative complications and late postoperative complications.

|  | **EXP group n (%)** | **TLE group n (%)** | **P-values†** |
| --- | --- | --- | --- |
| Early postoperative complication |  |  |  |
| Hyphema | 4 (13.3) | 2 (4.7) | 0.18 |
| Shallow anterior chamber | 5 (16.7) | 6 (14.0) | 0.70 |
| Choroidal detachment | 6 (20.0) | 4 (9.3) | 0.19 |
| Conjunctiva leakage | 5 (16.7) | 4 (9.3) | 0.35 |
| Hypotony maculopathy | 0 | 1 (2.3) | 0.40 |
| Total number of complications | 13 (43.3) | 10 (23.3) | 0.07 |
| Late postoperative complications |  |  |  |
| Hyphema | 0 | 0 | - |
| Shallow anterior chamber | 0 | 0 | - |
| Choroidal detachment | 0 | 1 (2.3) | 0.40 |
| Conjunctiva leakage | 0 | 1 (2.3) | 0.40 |
| Hypotony maculopathy | 0 | 0 | - |
| Total number of complications | 0 | 2 (4.7) | 0.27 |

†Pearson chi-square tests

### Risk factors for surgical failure by multivariate Cox proportional hazards model

The baseline characteristics, including history of cataract surgery, glaucoma diagnosis, preoperative IOP, and number of glaucoma medications were evaluated as prognostic factors for surgical failure (Table 4). The number of preoperative medications was a risk factor for failure to satisfy criteria A and B in the TLE group. Glaucoma surgery with cataract surgery reduced the risk of failure for criterion C in the EXP group. Factors not significantly associated with target IOP success rates included preoperative IOP, lens status, and diagnosis.

**Table 4.**
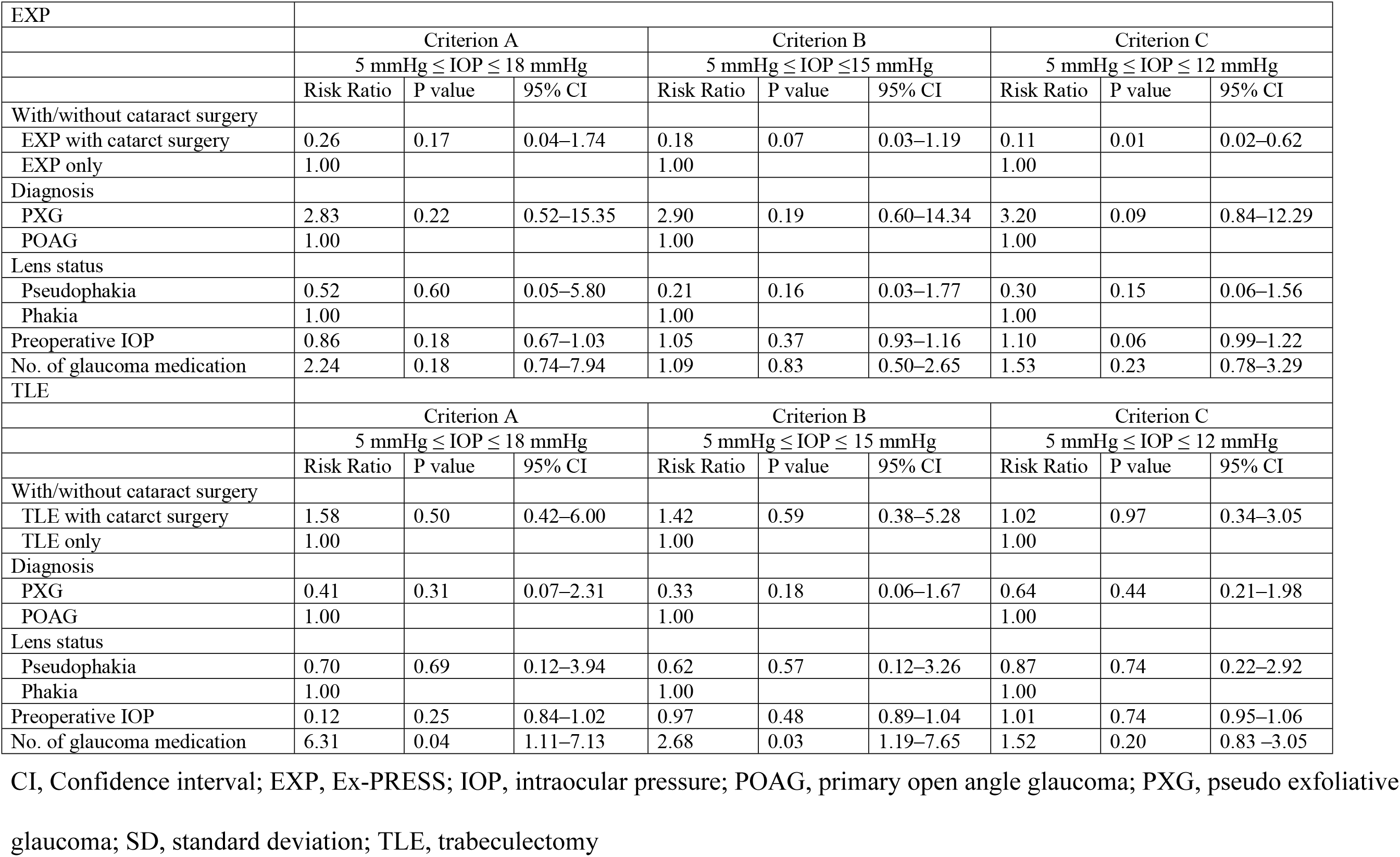
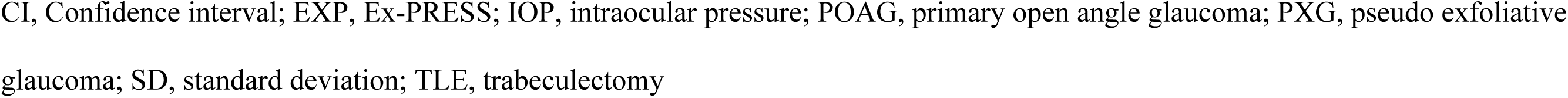
Multivariate Cox proportional hazard ratios for risk factors for failure.

## Discussion

Most previous studies defined successful TLE or EXP as a postoperative IOP of 18 mmHg or less. However, the Advanced Glaucoma Intervention Study reported that visual field impairment progressed unless the IOP was maintained at < 12.3 mmHg [8], and subjects with NTG often have IOP values < 18 mmHg. Therefore, in this study, we set the target IOP at several lower levels (criteria B and C).

Filtering surgery, including TLE, is a common and standard modality for patients with long-standing glaucoma. While we expected EXP to reduce IOP as effectively as TLE, based on previous studies [3-4,12], an equivalent efficacy has not been demonstrated for lower target IOP ranges. As the baseline IOP of some patients was in the normal range under maximum medical therapy, some studies have used the percentage reduction from baseline as an outcome metric in addition to absolute IOP levels. However, this criterion is not applicable in all situations. In the Japanese population, NTG accounts for the majority of glaucoma cases [10]. Low preoperative IOP as in NTG makes it difficult to achieve a 20% reduction from baseline after filtering surgery without complications [13-14]. Therefore, only patients with IOP ≥ 15 mmHg were included in this study.

Several previous prospective studies have compared the success rate of TLE to EXP both with and without the use of glaucoma medications. Gonzalez-Rodriguez et al [4] defined surgical success as 5 ≤IOP ≤18 (criterion A in our study) and a 20% reduction from baseline, and reported success rates of 59% and 76% for EXP and TLE groups, respectively, at 24 months and 52% and 61%, respectively, at 36 months. In accord with our results using criterion A, these rates did not differ between procedures.

Most previous studies have found no difference in the number of glaucoma medications required by patients following EXP or TLE [4,5,15-17], while a few have reported that the EXP group required fewer glaucoma medications [3,18] and one that patients required more medications following Ex-PRESS implantation [19]. In the current study, we found no significant difference in the number of glaucoma medications required before or after surgery. However, the EXP group required more glaucoma medications than the TLE group post surgery, and the group difference in the number of eye drops required increased gradually during follow-up. Thus, it is likely that the EXP group will eventually require a significantly greater number or significantly higher doses of glaucoma medications.

We found little difference in complication rates following EXP or TLE. One patient in the TLE group developed cataract, which resulted in loss of VA during the observation period, while there were no cases of cataract progression in the EXP group. Similarly, Beltran-Agullo and colleagues reported that cataract progression was more frequent following TLE than EXP [20], potentially because EXP does not require peripheral iridectomy and thus carries a lower risk of hypotony. Aqueous flare elevation may occur during or following filtration surgery due to disruption of the blood–aqueous barrier, and flare severity increases with inflammatory activity in the anterior segment [21]. One case study reported substantial flare on the first day after EXP that subsided by the third day [22], while another reported elevated flare at 4 weeks after TLE [23]. The more sustained flare after TLE may also result from iridectomy. Further, the reduced inflammation following EXP may prevent cataract progression. Moreover, Arimura and colleagues [16] reported a significantly greater total number of complications following TLE than following EXP. However, other studies have reported similar incidences of most early complications with the exception of hyphema [24]. In the current study as well, there were no significant differences in early and late postoperative complications between treatment groups. No late complications due to hypotony, such as hypotony maculopathy and choroidal detachment, were observed in the EXP group. However, one case in the TLE group exhibited choroidal detachment related to over filtration and low IOP. We assume that this is because EXP surgery creates a constant aqueous outflow route. From this, we expect that EXP is more suitable for patients at risk for complications related to low IOP. For example, young patients with long axial length tend to develop hypotony maculopathy after TLE [25], which could lead to a reduction in VA.

A Cox regression model identified the number of glaucoma medications before surgery as a risk factor for surgical failure in the TLE group, consistent with previous studies [26-27] and suggesting that preservatives of glaucoma medication increase the risk of TLE failure [28]. Postoperative subconjunctival fibrosis after ophthalmic surgery results in increased numbers of conjunctival fibroblasts and inflammatory cells [29], and preservatives will further increase the number of conjunctival fibroblasts and macrophages while reducing the number of conjunctival goblet cells [28]. These alterations lead to production of predisposition to heal the fistula drainage, resulting in uncontrolled IOP [26,28]. Since the role of the conjunctiva is the same in TLE and EXP, it can be expected that glaucoma medication will reduce the surgical success of EXP through similar mechanisms. However, the number of glaucoma medications was not a risk factor for EXP failure in this study. EXP does not require iridectomy and leads to less bleeding and inflammatory cell infiltration. As a result, fibrosis of the conjunctiva may be prevented after EXP, reducing the risk of surgical failure.

One limitation of this study is the group imbalance and relatively small number of patients because we exclude patients with preoperative IOP < 15 mmHg after randomization. We followed patients for 3 years in this study. The number of dropout patients increased with long-term follow-up, which is another limitation of this study. As glaucoma requires lifelong treatment, longer-term clinical follow is required.

In conclusion, no differences in surgical success, IOP, VA, and failure rates were noted between the EXP and TLE groups in this study. We also found that the number of glaucoma medications was a risk factor for surgical failure in the TLE group but not the EXP group.

## Data Availability

The research proposal stipulates that the data will be stored on a computer that is not connected to the Internet and will not be taken out.Data available on request from the author. The aurthor will provide data after reporting to the Ethics Committee of Hiroshima University Hospital.

## Acknowledgments

The authors would like to thank Enago (www.enago.jp) for the English language review

## Notes

### Competing Interest Statement

The authors have declared no competing interest.

### Clinical Trial

identifier University Hospital Medical Information Network UMIN000008981 date of access and registration, September 25, 2012

### Clinical Protocols

https://www.umin.ac.jp/english/

### Funding Statement

The author(s) received no specific funding for this work.

### Author Declarations

The institutional review board of Hiroshima University

